# Determinants of Severity in Cancer Patients with COVID-19 Illness

**DOI:** 10.1101/2020.05.04.20086322

**Authors:** Elizabeth V. Robilotti, N. Esther Babady, Peter A. Mead, Thierry Rolling, Rocio Perez-Johnston, Marilia Bernardes, Yael Bogler, Mario Caldararo, Cesar J. Figueroa Ortiz, Michael S. Glickman, Alexa Joanow, Anna Kaltsas, Yeon Joo Lee, Anabella Lucca Bianchi, Amanda Mariano, Sejal Morjaria, Tamara Nawar, Genovefa A. Papanicolaou, Jacqueline Predmore, Gil Redelman-Sidi, Elizabeth Schmidt, Susan K. Seo, Kent Sepkowitz, Monika Shah, Jedd D. Wolchok, Tobias M. Hohl, Ying Taur, Mini Kamboj

**Affiliations:** Infectious Diseases, Department of Medicine, Memorial Sloan Kettering Cancer Center, New York, NY; Infection Control, Department of Medicine, Memorial Sloan Kettering Cancer Center, New York, NY; Clinical Microbiology Service, Department of Laboratory Medicine, Memorial Sloan Kettering Cancer Center, New York, NY; Department of Radiology, Memorial Sloan Kettering Cancer Center, New York, NY; Employee Health Service, Memorial Sloan Kettering Cancer Center, New York, NY; Department of Medicine, Joan and Sanford Weill Medical College of Cornell University, New York, NY; Human Oncology and Pathogenesis Program, Department of Medicine, Ludwig Center and Parker Institute for Cancer Immunotherapy at Memorial Sloan Kettering Cancer Center, New York, NY

## Abstract

New York State had 180,458 cases of SARS-CoV-2 and 9385 reported deaths as of April 10^th^, 2020. Patients with cancer comprised 8.4% of deceased individuals^1^. Population-based studies from China and Italy suggested a higher COVID-19 death rate in patients with cancer^2,3^, although there is a knowledge gap as to which aspects of cancer and its treatment confer risk of severe COVID-19 disease^4^. This information is critical to balance the competing safety considerations of reducing SARS-CoV-2 exposure and cancer treatment continuation. Since March 10^th^, 2020 Memorial Sloan Kettering Cancer Center (MSKCC) performed diagnostic testing for SARS-CoV-2 in symptomatic patients. Overall, 40% out of 423 patients with cancer were hospitalized for COVID-19 illness, 20% developed severe respiratory illness, including 9% that required mechanical ventilation, and 9% that died. On multivariate analysis, age ≥ 65 years and treatment with immune checkpoint inhibitors (ICI) within 90 days were predictors for hospitalization and severe disease, while receipt of chemotherapy within 30 days and major surgery were not. Overall, COVID-19 illness is associated with higher rates of hospitalization and severe outcomes in patients with cancer. Association between ICI and COVID-19 outcomes will need interrogation in tumor-specific cohorts.

## MAIN

The characterization of COVID-19 in patients with cancer remains limited in published studies and nationwide surveillance analyses. Reports from outside the US show a higher risk of COVID-19 related severe events in patients on active therapy, those with lung cancer and ICI recipients^4–6^. In this study, we report on the epidemiology of COVID-19 illness experienced at our cancer center over the last month, during the height of incident cases in New York City, and offer an analysis of risk factors for severe infection that is pertinent to cancer patient populations. From March 10, 2020, until April 7, 2020, SARS-CoV-2 was detected in 530 (26%) of 2035 patients tested **(Figure 1)**. The final study population included 423 patients, with the following exclusions: outpatients with missing information (n = 77) and asymptomatic individuals tested as part of diagnostic screening protocols before surgery or myeloablative chemotherapy (n = 30). The MSKCC Institutional Review Board granted a HIPAA waiver of authorization to conduct the study.

**Figure 1:**
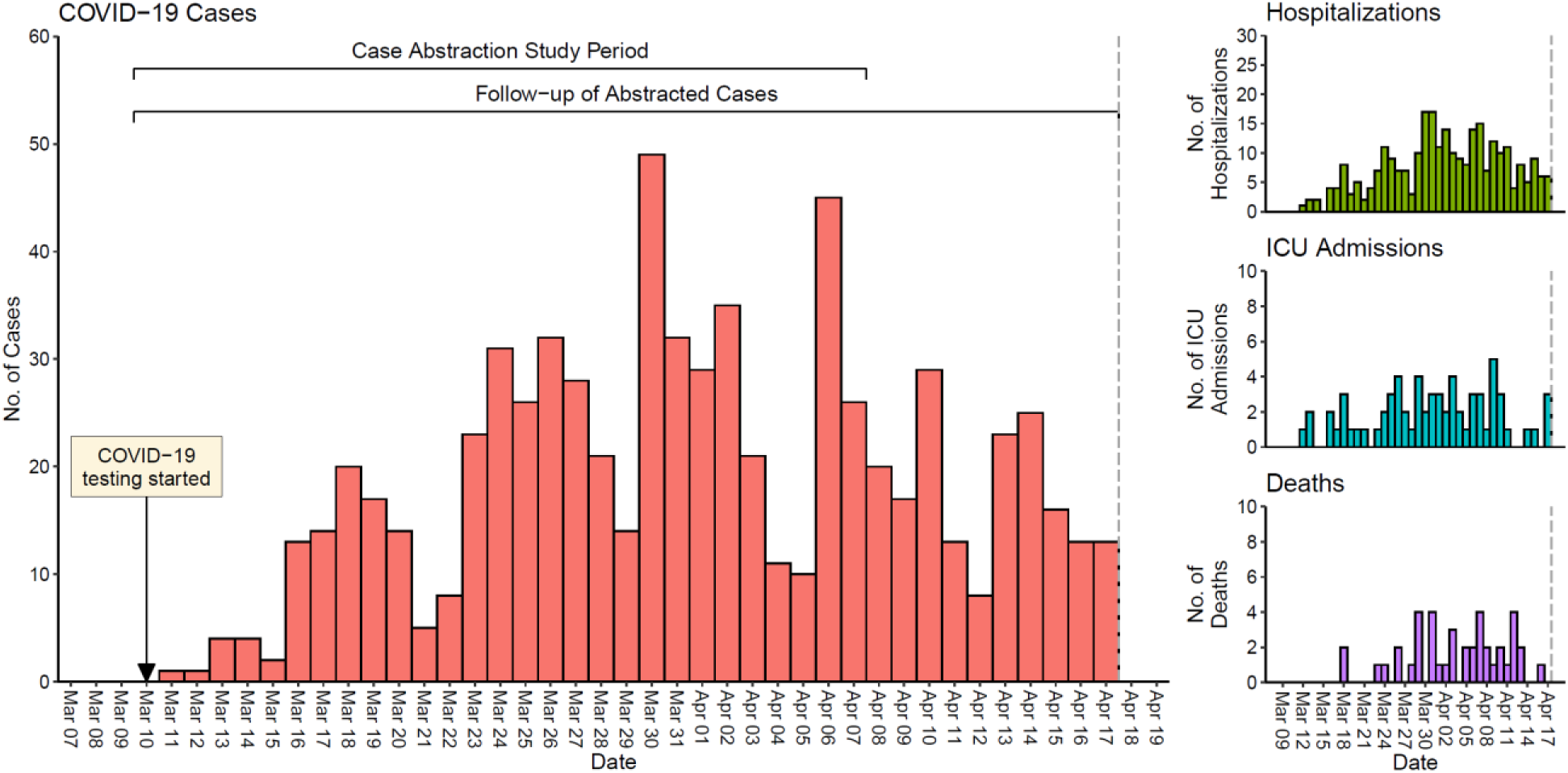
Number of SARS CoV-2 positive cases, hospitalizations, ICU admissions and deaths from March 10, 2020 to April 17, 2020

**Table 1** shows the demographic and clinical characteristics of the 423 cases. Most patients were adults over the age of 60 years (234, 55.3%). The most frequent cancer types included solid tumors such as breast (86, 20%), colorectal (37, 9%) and, lung cancer (35, 8%). Lymphoma was the most common hematologic malignancy (49, 12%). Over half of the cases were metastatic solid tumors (238, 56%). At least one of the specified co-morbid conditions were present in 234 (55.3%) individuals. The most frequent among these included hypertension, diabetes, and cardiovascular disease. Of the presenting symptoms we examined, fever (78%) and cough (82%) were the most common, whereas shortness of breath (44%) and diarrhea (26%) were less common but not rare. Chest radiographic findings are summarized in **Supplemental Table 1**. In the cohort, 168 (40%) out of 423 patients were hospitalized, and 84 (20%) developed severe respiratory illness, including 39 (9%) that required invasive mechanical ventilation. Illness in seven pediatric cases was mild and without complications. The overall case-fatality rate (CFR) was 9% (39 out of 423). Death rates for hospital and ICU admittance were 23% (39 of 168) and 33% (16 of 48), respectively.

**Table 1:** Patient demographics and clinical characteristics (N=423)

| **Characteristic** | **No. (%)** |
| --- | --- |
| **Age (years)** |  |
| <18 | 7 (2) |
| 18-29 | 11 (3) |
| 30-39 | 19 (4) |
| 40-49 | 51 (12) |
| 50-59 | 101 (24) |
| 60-69 | 134 (32) |
| ≥70 | 100 (24) |
| **Sex** |  |
| Male | 212 (50) |
| Female | 211 (50) |
| **Race** |  |
| White | 263 (62) |
| Black | 68 (16) |
| Asian | 36 (9) |
| Other | 56 (13) |
| **Body mass index (BMI)** |  |
| Below 18.5 (underweight) | 13 (3) |
| **18.5 to 24.9 (normal)** | 131 (31) |
| **25.0 to 29.9 (overweight)** | 149 (35) |
| **30.0 to 39.9 (obesity)** | 111 (26) |
| **40 or higher (severe obesity)** | 19 (4) |
| **Smoking status** |  |
| **Current** | 9 (2) |
| **Former** | 159 (38) |
| **Never** | 248 (59) |
| **Unknown** | 7 (2) |
| **Underlying Cancer** |  |
| **Hematologic** |  |
| **Leukemia** | 32 (8) |
| **Lymphoma** | 49 (12) |
| **Myeloma** | 22 (5) |
| **Solid Tumor** |  |
| **Breast** | 86 (20) |
| **Colorectal** | 37 (9) |
| **Lung** | 35 (8) |
| **Prostate** | 26 (6) |
| **Other** | 136 (32) |
| **Metastatic disease** | 238 (56) |
| **Major surgery (30d)** | 31 (7) |
| **Asthma** | 43 (10) |
| **COPD** | 29 (7) |
| **Diabetes** | 84 (20) |
| **Cardiac dysfunction** | 84 (20) |
| **Chronic kidney disease** | 36 (9) |
| **Hypertension** | 214 (51) |
| **Systemic chemotherapy (30d)** | 191 (45) |
| **Chronic corticosteroid** | 66 (16) |
| **Chronic lymphopenia** | 9 (2) |
| **Immune checkpoint inhibitor (ICI)<sup>a</sup>** | 31 (7) |
| **Symptoms at Onset** |  |
| **Fever** | 331 (78) |
| **Shortness of breath** | 186 (44) |
| **Cough** | 346 (82) |
| **Diarrhea** | 108 (26) |
| **Outcomes (30d)** |  |
| **Hospitalization status** |  |
| **Not admitted** | 243 (57) |
| **Admitted** | 168 (40) |
| **Already hospitalized** | 12 (3) |
| **Oxygen requirements<sup>b</sup>** |  |
| **None<sup>c</sup>** | 287 (68) |
| **Low-flow oxygen** | 52 (12) |
| **High-flow oxygen** | 45 (11) |
| **Mechanical ventilation** | 39 (9) |
| **Death** | 39 (9) |

<sup>a</sup> Includes 24 patients on PD-1 and five patients on PD-L1 inhibitors.
<sup>b</sup> Highest oxygen requirement is shown.
<sup>c</sup> Note, 2 patients had chronic tracheostomy because of prior medical issues, but did not require supplemental oxygen during COVID-19 infection.

Next, we assessed risk factors for hospitalization and severe respiratory illness, defined as the requirement for high-flow oxygen supplementation or mechanical ventilation. In the multivariate analysis, the following risk factors were independently associated with hospitalization: age > 65 years, non-white race, hematologic malignancy, a composite measure of chronic lymphopenia and/or corticosteroid use, and treatment with ICI therapy. Hypertension and/or chronic kidney disease trended towards significance as predictors of hospitalization in the multivariate model **(Table 2)**. The risk factors for severe respiratory illness due to COVID-19 were overlapping with those for hospitalization, but not identical. Severe illness was significantly more common with age > 65 years. Of note, treatment with ICI also remained an independent predictor of severe respiratory illness. Metastatic disease, recent receipt of chemotherapy, or major surgery within the previous 30 days did not show a significant association with either hospitalization or severe respiratory illness **(Table 3)**. Given the apparent association of ICI with COVID-19 severity, we explored this further by calculating stratum-specific rates of hospitalization and severe respiratory illness **(Table 4)**. Since PD-1, a type of ICI, is commonly used to treat lung cancer, we examined the occurrence of outcomes by ICI use and underlying cancer (lung vs non-lung). The frequency of severe illness and hospitalization was higher in both ICI treated groups. In a separate prediction model, clinical presentation with new onset dyspnea and diarrhea were most predictive of hospitalization and severe illness **(Supplemental Table 2)**.

**Table 2:** Predictors of hospitalization for COVID-19, by logistic regression (N=411^a^)

| *Predictor* | *Univariate* |  | *Multivariate* |  |
| --- | --- | --- | --- | --- |
|  | *OR (95% CI)* | *P-value* | *OR (95% CI)* | *P-value* |
| *Age (>65)* | 1.81 (1.20 - 2.72) | 0.004 | 1.58 (1.00 - 2.50) | 0.05 |
| *Sex (female)* | 0.89 (0.60 - 1.32) | 0.58 |  |  |
| *Race (non-white)* | 1.36 (0.91 - 2.04) | 0.14 | 1.59 (1.04 - 2.46) | 0.03 |
| *BMI (≥30)* | 0.87 (0.57 - 1.33) | 0.53 |  |  |
| *Current Smoker* | 1.16 (0.28 - 4.45) | 0.83 |  |  |
| *Asthma/COPD* | 1.39 (0.81 - 2.37) | 0.27 | 1.14 (0.64 - 2.03) | 0.66 |
| *Cancer (non-met solid)* | 1.00 (Ref) | - | 1.00 (Ref) |  |
| *Cancer (met solid)* | 0.87 (0.52 - 1.48) | 0.60 | 0.74 (0.42-1.30) | 0.29 |
| *Cancer (hematologic)* | 2.15 (1.20 - 3.90) | 0.010 | 2.37 (1.28 - 4.43) | 0.006 |
| *Major Surgery (within 30d)* | 1.24 (0.53 - 2.84) | 0.61 |  |  |
| *Diabetes* | 1.20 (0.73 - 1.96) | 0.47 |  |  |
| *Cardiac Disorder<sup>b</sup>* | 1.86 (1.13 - 3.07) | 0.02 | 1.37 (0.79 - 2.40) | 0.26 |
| *HTN/chronic kidney disease* | 1.84 (1.24 - 2.75) | 0.003 | 1.53 (0.97 - 2.42) | 0.07 |
| *Chemotherapy<sup>c</sup> (last 30d)* | 1.04 (0.70 - 1.54) | 0.85 |  |  |
| *Chronic lymphopenia<sup>d</sup> or*<br>*Corticosteroids<sup>e</sup>* | 1.86 (1.11 - 3.15) | 0.02 | 1.85 (1.06 - 3.22) | 0.03 |
| *Immune checkpoint inhibitor<sup>f</sup>* | 2.53 (1.18 - 5.67) | 0.02 | 3.06 (1.35 - 7.20) | 0.007 |

COPD chronic obstructive pulmonary disease; HTN hypertension
<sup>a</sup> 12 subjects were excluded from this analysis; they were already hospitalized for other reasons, prior to developing COVID-19 infection.
<sup>b</sup> Heart failure, myocardial infarction, valvular replacement, or cardiomyopathy
<sup>c</sup> Systemic, parenteral chemotherapy
<sup>d</sup> Absolute lymphocyte count <500 per microliter over five previous consecutive measurements
<sup>e</sup> Corticosteroids (equivalent of prednisone 20 mg or higher) for at least 10 days.
<sup>f</sup> Any of the following within 90 days: ipilimumab, pembrolizumab, nivolumab, atezolizumab, avelumab, durvalumab, tremelimumab

**Table 3:** Predictors of severe respiratory illness due to COVID-19, by Cox Proportional Hazard (N=423)

| *Predictor* | *Univariate* |  | *Multivariate* |  |
| --- | --- | --- | --- | --- |
|  | *HR (95% CI)* | *P-value* | *HR (95% CI)* | *P-value* |
| *Age (>65)* | 2.18 (1.42 - 3.34) | <0.001 | 1.80 (1.14 - 2.84) | 0.01 |
| *Sex (female)* | 1.07 (0.70 - 1.64) | 0.75 |  |  |
| *Race (non-white)* | 1.17 (0.76 - 1.81) | 0.49 |  |  |
| *BMI (≥30)* | 1.05 (0.67 - 1.67) | 0.82 |  |  |
| *Smoking* | 1.12 (0.28 - 4.56) | 0.87 |  |  |
| *Asthma/COPD* | 1.70 (1.02 - 2.84) | 0.04 | 1.39 (0.82 - 2.36) | 0.22 |
| *Cancer (non-met solid)* | 1.00 (Ref) | - | 1.00 (Ref) |  |
| *Cancer (met solid)* | 0.83 (0.46-1.51) | 0.55 | 0.69 (0.37-1.31) | 0.26 |
| *Cancer (hematologic)* | 1.53 (0.83 - 2.83) | 0.17 | 1.61 (0.86 - 2.99) | 0.14 |
| *Major Surgery (within 30d)* | 1.34 (0.65 - 2.78) | 0.43 |  |  |
| *Diabetes* | 1.15 (0.68 - 1.94) | 0.60 |  |  |
| *Cardiac Disorder<sup>a</sup>* | 2.12 (1.34 - 3.35) | 0.001 | 1.48 (0.89 - 2.44) | 0.13 |
| *HTN/chronic kidney disease* | 1.83 (1.17 - 2.86) | 0.008 | 1.27 (0.78 - 2.08) | 0.34 |
| *Chemotherapy<sup>b</sup> (last 30d)* | 1.16 (0.76 - 1.78) | 0.49 |  |  |
| *Chronic lymphopenic<sup>3</sup> or corticosteroids<sup>d</sup>* | 1.67 (1.02 - 2.74) | 0.04 | 1.52 (0.92 - 2.50) | 0.10 |
| *Immune checkpoint inhibitor<sup>e</sup>* | 2.45 (1.33 - 4.51) | 0.004 | 3.03 (1.53 - 5.98) | 0.001 |

COPD chronic obstructive pulmonary disease; HTN hypertension
<sup>a</sup> Heart failure, myocardial infarction, valvular replacement, or cardiomyopathy
<sup>b</sup> Systemic, parenteral chemotherapy
<sup>c</sup> Absolute lymphocyte count <500 per microliter over 5 previous consecutive measurements
<sup>d</sup> Corticosteroids (equivalent of prednisone 20 mg or higher) for at least 10 days.
<sup>e</sup> Any of the following within 90 days: ipilimumab, pembrolizumab, nivolumab, atezolizumab, avelumab, durvalumab, tremelimumab

**Table 4:** Stratum-specific point estimates of outcomes, by immune checkpoint inhibitor (ICI) treatment and underlying solid cancer (lung vs. non-lung) (N=226)

| *Cancer* | *Endpoint* | *Non-ICI no./total no. (%)* | *ICI<sup>c</sup> no./total no. (%)* |
| --- | --- | --- | --- |
| *Lung cancer—* | Hospitalization<sup>b</sup> | 12/23 (52) | 10/12 (83) |
|  | Severe respiratory illness | 8/23 (35) | 7/12 (58) |
| *Other solid cancers<sup>a</sup>* | Hospitalization<sup>b</sup> | 82/216 (38) | 8/17 (47) |
|  | Severe respiratory illness | 34/221 (15) | 5/19 (26) |

<sup>a</sup> Includes the following malignancies where ICI was given as therapy: breast, colorectal, genitourinary, gynecologic, lymphoma, neurologic, skin.
<sup>b</sup> Exclusive of patients already admitted to the hospital at time of COVID-19 diagnosis
<sup>c</sup> Within 90 days

Patients with cancer are among those most vulnerable to severe illness from respiratory viral infections.^7^ Our early experience with COVID-19 at a large tertiary care cancer center demonstrated severe disease in 20% of patients diagnosed with COVID-19 illness, with an overall CFR of 9%. Similar to other studies in the general population, we found that age, non- white race, cardiac disease, hypertension, and chronic kidney disease correlated with severe outcomes.^8,9^ Contrary to previous reports, receipt of chemotherapy within 30 days before COVID-19 diagnosis was not associated with a higher risk of complications.^6^ Recent major surgery and metastatic disease also did not confer a significant risk of severe COVID-19 disease. Treatment with ICI predicted both hospitalization and severe disease, although there was considerable heterogeneity in ICI treated tumor types, and disease-specific confounding factors could not be specifically addressed. COVID-19 illness among children with cancer exhibited a milder course, consistent with reports in children without cancer, but represented a small portion of the evaluated population (7, 2%).^10^

Immunotherapy was an independent risk factor in our study involving 31 ICI-treated patients, 12 of 31 had lung cancer, while the remaining reflected a combination of multiple primary tumor groups. While we observed more severe COVID-19 illness in ICI recipients with underlying lung cancer, non-lung cancer patients treated with ICI also demonstrated severe outcomes **(Table 4)**. A possible explanation for this observation is an exacerbation of ICI-related lung injury, or ICI-triggered immune dysregulation by T cell hyperactivation, that in turn may facilitate acute respiratory distress syndrome, a dreaded COVID-19 disease complication.^11^ This finding should be interpreted with caution due to the limitations of our ICI dataset. It is possible that patients with lung cancer have confounding from other risk factors beyond ICI treatment (e.g., surgery, preexisting lung disease, radiotherapy, prior smoking) that contributed to the finding of increased disease severity and were not fully evaluable in our population. Until more extensive studies are available, it is prudent not to alter treatment decisions, but consider SARS CoV-2 testing for patients initiating or continuing treatment with ICIs irrespective of symptoms.

There are several other limitations to our study. First, we describe a single center retrospective analysis in a heterogeneous group of patients with cancer. Second, outcomes may have been incomplete, given the short follow-up period and the possibility of reporting delays; 49 patients remain hospitalized at the time of this report. Third, many patients received an array of medications, including hydroxychloroquine, remdesivir, corticosteroids, and IL-6 inhibitors, according to the decision of the primary team caring for the patient. The effectiveness of these experimental therapeutics was not explicitly evaluated. Finally, with the postponement of all non-essential cancer care during the study period, COVID-19 testing practices were mostly targeted towards symptomatic patients that needed medical evaluation, potentially overestimating the overall severity of COVID-19.

In summary, the outcome of COVID-19 illness is worse among those with underlying conditions, including cancer. Our group of 423 patients with cancer had substantially higher rates of severe outcomes with COVID-19 illness when compared to published worldwide rates of severe disease (6.1 -31.4%) and death (2.3-7.2%).^3,12–14^ In addition, as was seen with the SARS epidemic in 2003, the ongoing risk of contracting the illness and indirect consequences of treatment disruptions are expected to have a lasting effect on the health and safety of patients undergoing treatment for cancer.^15^ Continuous preparedness is paramount as routine cancer care is resumed in the coming weeks and months amidst the unpredictable threat posed by COVID-19. Informed approaches with universal screening, aggressive testing, and rigorous control measures will be essential for the safe ongoing delivery of oncologic care.

## Data Availability

All data reported in the manuscript will be made available upon request from the corresponding author following editorial acceptance in a peer-reviewed journal.

## Acknowledgements

Supported by the Memorial Sloan Kettering Cancer Center core grant (P30 CA008748), the Burroughs Wellcome Fund Investigator in the Pathogenesis of Infectious Diseases Award (TMH), the Ludwig Collaborative and Swim Across America Laboratory (JDW), the Parker Institute for Cancer Immunotherapy (JDW), Department of Medicine, Memorial Sloan Kettering Cancer Center and Weill Cornell Medicine.

## Author Contribution

EVR, MK, YT, MSG made substantial contribution to study conception and design. YT, EVR, PAM, TR, MK acquisition of data. YT and TR conducted statistical analysis. TMH, MSG, MK, YT, EVR interpretation of data. MK drafted the first version of the article. All authors contributed to critical revisions and approved the final version. MK, YT, TMH, EVR take responsibility for the integrity of the work.

## Competing interests

The authors declare no competing interests.

**Supplemental Table 1:** Chest Radiographic Findings in COVID-19 cases

| **Chest radiograph (X-ray) findings** | **No. (%)** |
| --- | --- |
| **Normal** | 58 (28) |
| **Abnormal** | 149 (72) |
| **Total performed** | 207 (100) |

| **Computed tomography (CT) chest findings** | **No. (%)** |
| --- | --- |
| **Normal** | 4 (10) |
| **Abnormal** | 35 (90) |
| **Unilateral** | 12 (31) |
| **Bilateral** | 23 (59) |
| **Peripheral involvement** | 30 (77) |
| **Ground-glass infiltrates** | 30 (77) |
| **Focal consolidation** | 13 (33) |
| **“Crazy paving” finding** | 2 (5) |
| **Pleural effusion** | 5 (13) |
| **Adenopathy** | 1 (3) |
| **Total performed** | 39 (100) |

**Supplemental Table 2:** Presenting symptoms as predictors of subsequent hospitalization and severe respiratory illness for patients with COVID-19

**Hospitalization, by logistic regression (N=411)**
| *Predictor* | *Univariate* |  | *Multivariate* |  |
| --- | --- | --- | --- | --- |
|  | *OR (95% CI)* | *P-value* | *OR (95% CI)* | *P-value* |
| *Fever* | 1.79 (1.10 - 2.98) | 0.020 | 1.44 (0.86 - 2.48) | 0.170 |
| *Cough* | 2.13 (1.25 - 3.78) | 0.005 | 1.54 (0.86 - 2.82) | 0.149 |
| *Shortness of breath* | 3.58 (2.39 - 5.41) | <0.001 | 3.05 (2.00 - 4.68) | <0.001 |
| *Diarrhea* | 2.66 (1.70 - 4.17) | <0.001 | 2.32 (1.45 - 3.73) | <0.001 |

**Severe respiratory illness, by Cox proportional hazard (N=423)**
| *Predictor* | *Univariate* |  | *Multivariate* |  |
| --- | --- | --- | --- | --- |
|  | *HR (95% CI)* | *P-value* | *HR (95% CI)* | *P-value* |
| *Fever* | 1.53 (0.85 - 2.76) | 0.159 | 1.17 (0.64 - 2.13) | 0.613 |
| *Cough* | 1.71 (0.88 - 3.31) | 0.112 | 1.11 (0.56 - 2.19) | 0.766 |
| *Shortness of breath* | 4.88 (2.95 - 8.07) | <0.001 | 4.42 (2.64 - 7.41) | <0.001 |
| *Diarrhea* | 2.27 (1.47 - 3.51) | <0.001 | 1.89 (1.22 - 2.94) | 0.004 |

## ONLINE METHODS

Memorial Sloan Kettering Cancer Center (MSKCC) is a 514-bed tertiary cancer center in New York City with approximately 25,000 admissions and 173,000 patient days annually. MSKCC maintains 19 ambulatory sites across New York State and New Jersey, with in excess of a million combined yearly outpatient visits. Approximately 23,000 individuals per year are under active treatment. During February and March 2020, a total of 14,067 individuals received parenteral chemotherapy at MSKCC.

### Study population

From March 10, 2020, until April 7, 2020, all consecutive adult and pediatric cases of symptomatic and laboratory-confirmed SARS-CoV-2 infection were included. The only exceptions were asymptomatic subjects tested before surgery or prior to receipt of select myeloablative chemotherapy regimens. Identification of case-patients, their medical background, and clinical course during COVID-19 illness, were abstracted from electronic medical records.

### Laboratory methods

Nasopharyngeal swab samples were collected using flocked swabs (Copan Diagnostics) and placed in viral transport media. SARS-CoV-2 RNA was detected using the Centers for Disease Control and Prevention (CDC) protocol targeting two regions of the nucleocapsid gene (N1 and N2), with the following modifications: Nucleic acids were extracted from specimens using the NUCLISENS EasyMag (bioMérieux, Durham, NC) following an off-board, pre-lysis step.^16,17^ Real-time reverse transcription PCR was performed on the ABI 7500 Fast (Applied Biosystems Foster City, CA) in a final reaction volume of 20-μL, including 5 μL of extracted nucleic acids. Samples were reported as positive if both the N1 and N2 targets were detected.

### Statistical analysis

We first assessed patient risk factors for hospitalization as part of the management for COVID-19 illness, using logistic regression. Patients with nosocomial infection (n=12) were excluded from this analysis. Next, we assessed risk factors for severe respiratory illness, defined as the requirement for high-flow oxygen supplementation or mechanical ventilation. Cause-specific Cox proportional hazard modeling was applied for this. Analysis time began at the time of COVID-19 diagnosis and continued until one of the following endpoints was achieved: 30 days since diagnosis, the end of the study follow-up period (April 17, 2020), or death.

For the outcomes described above, the following clinical variables were assessed: age, sex, race, diabetes, hypertension cardiovascular disease (myocardial infarction, heart failure, heart valve replacement, or cardiomyopathy), chronic obstructive pulmonary disease, asthma, chronic kidney disease, obesity (body mass index >30), smoking status, underlying cancer, chronic lymphopenia (absolute lymphocyte count less than 500 per microliter for at least five measurements, immediately preceding positive COVID-19 PCR test), chronic corticosteroid use (prednisone of 20 mg per day or equivalent, for at least ten days), systemic parenteral chemotherapy within 30 days, and major surgery within 30 days, treatment with an immune checkpoint inhibitors (ICI) within 90 days. In addition to past underlying conditions, new symptoms at the time of testing were assessed: fever, cough, shortness of breath, and diarrhea.

For both outcomes, predictors were first analyzed separately in a univariate analysis. Predictors with a univariate *p*-value of less than 0.25 were incorporated into a multivariate model. All study analyses were performed on R version 3.5 (R Development Core Team, Vienna, Austria). The MSKCC Institutional Review Board granted a HIPAA waiver of authorization to conduct the study.

## References

1. New York State Department of Health. COVID-19 Tracker. https://covid19tracker.health.ny.gov/views/NYS-COVID19-Tracker/NYSDOHCOVID-19Tracker-Fatalities (2020).

2. World Health Organization. Report of the WHO-China Joint Mission on Coronavirus Disease 2019 (COVID-19) (2020).

3. Onder, G., Rezza, G. & Brusaferro, S. Case-Fatality Rate and Characteristics of Patients Dying in Relation to COVID-19 in Italy. JAMA (2020).

4. Yu, J., Ouyang, W., Chua, M.L.K. & Xie, C. SARS-CoV-2 Transmission in Patients With Cancer at a Tertiary Care Hospital in Wuhan, China. JAMA Oncol (2020).

5. Liang, W., et al. Cancer patients in SARS-CoV-2 infection: a nationwide analysis in China. Lancet Oncol 21, 335–337 (2020).

6. Dai, M.-Y., Liu, D, Liu, M, et al Patients with Cancer Appear More Vulnerable to SARS-CoV-2: A Multi-Center Study During the COVID-19 Outbreak. Lancet (2020).

7. Hijano, D.R., Maron, G. & Hayden, R.T. Respiratory Viral Infections in Patients With Cancer or Undergoing Hematopoietic Cell Transplant. Front Microbiol 9, 3097 (2018).

8. Garg S K.L., et al. Hospitalization Rates and Characteristics of Patients Hospitalized with Laboratory-Confirmed Coronavirus Disease 2019 - COVID-NET, 14 States, March 1-30, 2020. MMWR Morb Mortal Wkly Rep 69, 458–464 (2020).

9. Centeres for Disease Control and Prevention COVID-19 Response Team. Preliminary Estimates of the Prevalence of Selected Underlying Health Conditions Among Patients with Coronavirus Disease 2019 — United States, February 12–March 28, 2020. MMWR Morb Mort Wkly Rep 69, 382–386 (2020).

10. Centeres for Disease Control and Prevention COVID-19 Response Team. Coronavirus Disease 2019 in Children - United States, February 12-April 2, 2020. MMWR Morb Mortal Wkly Rep 69, 422–426 (2020).

11. Xu, Z., et al. Pathological findings of COVID-19 associated with acute respiratory distress syndrome. Lancet Respir Med 8, 420–422 (2020).

12. Guan, W.J. & Zhong, N.S. Clinical Characteristics of Covid-19 in China. Reply. N Engl J Med 382(2020).

13. Centeres for Disease Control and Prevention COVID-19 Response Team. Severe Outcomes Among Patients with Coronavirus Disease 2019 (COVID-19)—United States, February 12–March 16, 2020. MMWR Morb Mortal Wkly Rep 69, 343–346 (2020).

14. Wu, Z. & McGoogan, J.M. Characteristics of and Important Lessons From the Coronavirus Disease 2019 (COVID-19) Outbreak in China: Summary of a Report of 72314 Cases From the Chinese Center for Disease Control and Prevention. JAMA (2020).

15. Hwang, S.W., Cheung, A.M., Moineddin, R. & Bell, C.M. Population mortality during the outbreak of Severe Acute Respiratory Syndrome in Toronto. BMC Public Health 7, 93 (2007).

16. Centeres for Disease Control and Prevention. Real-Time RT-PCR Panel for Detection 2019-Novel Coronavirus. https://www.cdc.gov/coronavirus/2019-ncov/downloads/rt-pcr-panel-for-detection-instructions.pdf (2020).

17. Centeres for Disease Control and Prevention. 2019-Novel Coronavirus (2019-nCoV) Real-time rRT-PCR Panel Primers and Probes. https://www.cdc.gov/coronavirus/2019-ncov/downloads/rt-pcr-panel-primer-probes.pdf (2020).

